# Adherence to iron and folic acid supplementation among pregnant women in rural areas of Eastern Tigray, Ethiopia: A community-based study

**DOI:** 10.64898/2026.11.18.26368382

**Authors:** Lemlem Asheber Alemu, Hadush Negash Meles, Tsige Shishay, Abadi Kidanemariam Berhe, Miglas Welay Gebregergis, Berhane Gebregziabiher, Yemane Berhane Tesfau

## Abstract

**Background:** Poor adherence to Iron and folic acid (IFA) remains major public health challenge in Ethiopia. While national protocols advise IFA supplementation during pregnancy, adherence is low in the nation particularly in rural areas. In the study setting, there is lack of updated data regarding adherence to IFA and its determinant factors among pregnant women.

**Objective:** To assess adherence to IFA supplements and associated factors among pregnant women in rural setting of Eastern Tigray, Ethiopia, 2025.

**Methods:** A community-based cross-sectional study was carried out from March 2025 to April 2025 among 614 pregnant women in Eastern Tigray, Ethiopia. Simple random and systematic random sampling techniques were utilized to select districts and study participants respectively. Data analysis was performed using SPSS 25, employing both binary and multivariable logistic regression. Multi-collinearity and goodness of fit tests were ensured using the variance inflation factor and Hosmer-Lemeshow test respectively. Statistical significance was declared at a p-value less than 0.05.

**Results:** A total of 599 participants were included and the prevalence of adherence to IFA Supplementation (IFAS) was 56% (95% CI: 52%, 60%). Having ≥ 4 ANC visits (AOR= 37.7 95% CI: 14.08 - 101.03), mothers with previous history of anemia (AOR=2.2 CI: 1.07 - 4.69), age of mothers ≥ 40 years (AOR= 11.5 CI: 1.06 - 124.05), mothers who received counseling about IFAS (AOR, 7.1; CI 3.24 - 15.37), and mothers travelled to the health facility 30 to 60 minutes and more than 60 minutes (AOR, 0.02; CI 0.01 - 0.04), and (AOR, 0.01(0 - 0.004), respectively had statistically significant association with adherence to IFAS.

**Conclusion:** Women having four or more ANC visits, previous history of anemia, receiving counseling about IFAS, elder age, and travelling less than 60 minutes to seek healthcare were associated with increased odds of adherence to IFAS.

## Introduction

Nutritional anemia is a major public health problem throughout the world, particularly in developing countries and evidence suggests that iron deficiency is the most common causative (1). Globally, 1.62 billion people suffer from anemia that accounts for 24.8% of the world’s population. In the past three decades, the global prevalence of anemia among pregnant women aged 15-49 years has dropped to 35.5% as of recent updates, though it remains a severe public health challenge developing nations (2). In Ethiopia, 31.7% (3–5) and 24% (6) of pregnant women and women of reproductive age are anemic respectively. This contributes to 18% of perinatal mortality, 19% preterm births, and 12% of low birth weight in low and middle-income countries. To combat this, daily intake of 30mg to 60mg of iron and 0.4mg folic acid is highly recommended for pregnant women (7).

The supplementation helps a growing fetus and pregnant women for enhanced DNA synthesis and cell division during early stages of fetal development to prevent neural tube defects (7). Consistent daily IFAS during pregnancy has been shown to reduce the overall risk of maternal anaemia by 70% and iron deficiency anemia (IDA) at term by 57% (7, 8). It prevents perinatal complications like pre-eclampsia, low birth weight, and prematurity (9, 10).

Despite these clinical benefits, adherence to IFAS remains low in Ethiopia (10, 11). The post-war in Tigray exacerbates the implementation gap, especially in rural areas where low health infrastructure, socioeconomic and geographic constraints, and other factors further hinder consistent supplement uptake.

Therefore, the study helps for a multifaceted approach to improve maternal health, decreases susceptibility to infection, decreases complications during childbirth, prevents neural tube defects, and helps to understand and address the factors affecting adherence to IFAS.

## Methods

### Study setting

The study was conducted in Eastern Tigray, Northern Ethiopia. Eastern Tigray comprises 7 urban districts, 11 rural districts and 156 kebele, and zonal administrative town. It is bounded in the east by Afar region, in the west by the Central Tigray, in the north by Eritrea and in the south by South-east Tigray. Eastern Tigray has two General Hospital, 5 Primary Hospital, 40 Health Centers and 145 Health Posts. According to the zonal office demographic data for 2023/2024, the zone has a total population of 1,051,177. Among them 536,100 are females and about 246,816 are reproductive age groups. There are 32,583 live births in the zone. The study was conducted from March to April 2025.

### Study design

A community-based cross-sectional study design was employed.

### Population

#### Source population

All pregnant women in rural Eastern zone of Tigray, Northern Ethiopia

#### Study population

All pregnant women in the selected Kebeles of rural Eastern zone, Tigray, northern Ethiopia

#### Study participants

All interviewed pregnant women in rural Eastern zone, Tigray, northern Ethiopia

### Eligibility criteria

#### Inclusion criteria

All pregnant women in the rural Eastern Zone of Tigray who attended antenatal care (ANC) for at least their second visit, and/or those who received iron and folic acid supplementation services from Health Extension Workers (HEWs) at home.

#### Exclusion criteria

Pregnant women who are critically ill at the time of data collection were excluded.

### Sampling technique and sample size determination

#### Sample size

The sample size for the first specific objective was calculated by using the single population proportion formula. The magnitude of adherence to iron foliate supplementation is taken 41.38% from a systematic review in Ethiopia (12) with a precision of 5% and 95% CI. Using design effect of 1.5 and 10% non-response rate, the total sample size is 614.

Applying the formula:

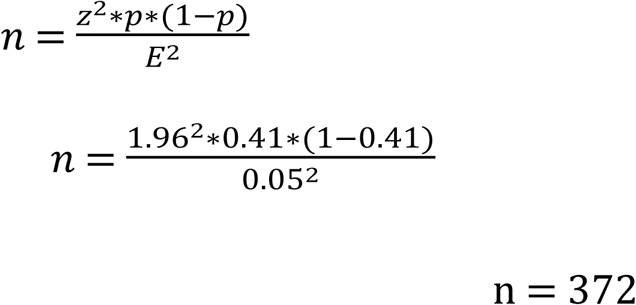

Considering design effect of 1.5 the sample size will be 558

By adding 10% non-response rate, the total required sample size will be n= 614

For the second specific objective sample size is calculated considering two population proportions using Epi-info version 7. To calculate sample size 95% level of confidence and 80% power were considered. Finally larger sample size (n = 614) was used in the study.

#### Sampling technique

Out of the 11 rural districts of the zone, 4 districts were selected using simple random sampling. Study participants were allocated proportionally from each kebeles based on population size of each kebeles from a list of households having pregnant women (**Figure 1**).

**Figure 1.**
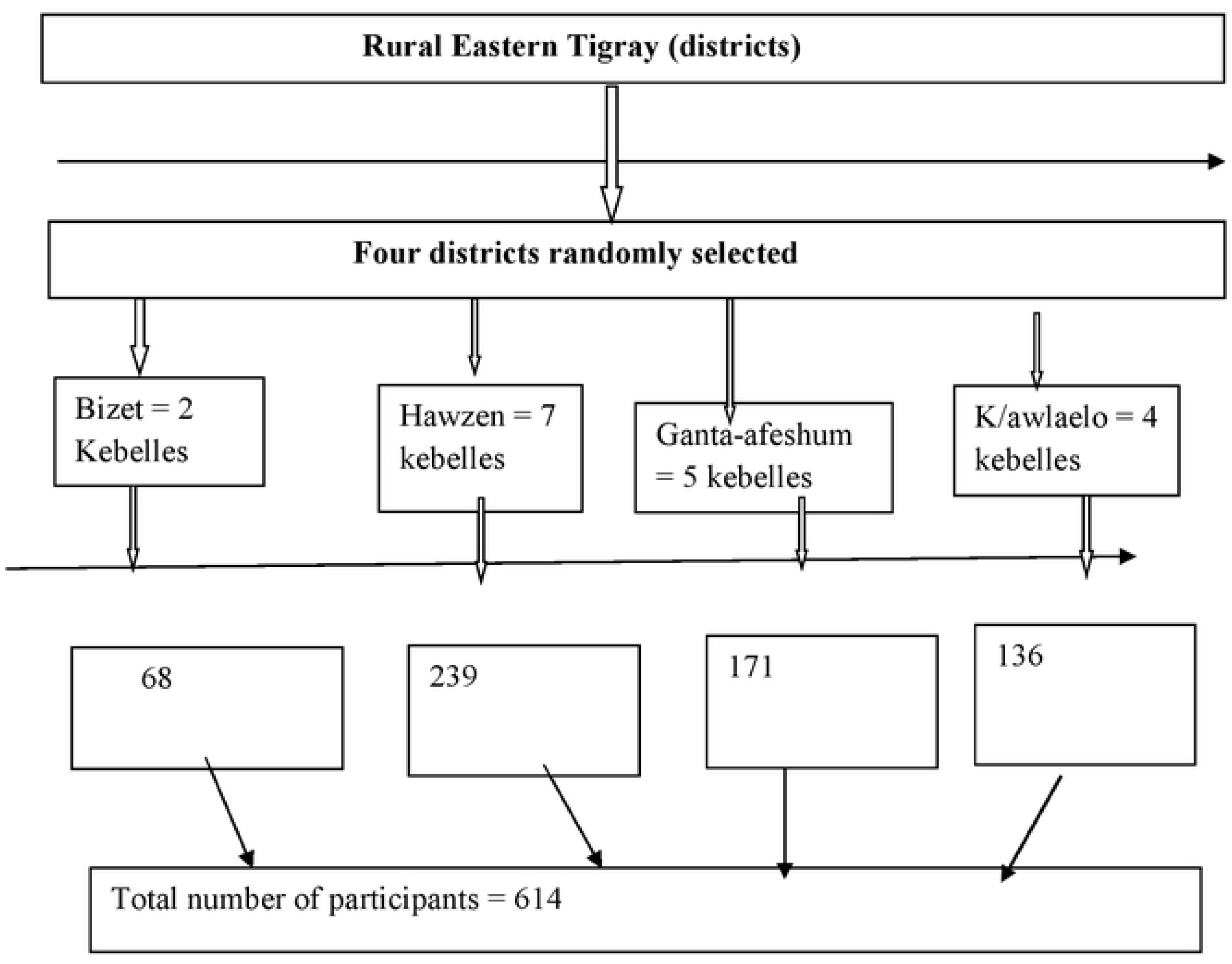
Schematic presentation of the sampling procedure of this study, Eastern Tigray, 2025.

### Study variables

#### Dependent variable

- Adherence to IFAS

#### Independent variables

- Socio-demographic characteristics like age, sex, marital status, educational background, income level, family size, religion, ethnicity and occupational status, obstetric and health-related characteristics of pregnant women attending ANC Clinic.

### Operational definitions

**Adherence to IFAS**: Pregnant women are said adherent to IFAS if they took the supplement at least 4 days a week in the previous month of IFAS (13).

**Good knowledge about IFAS**: Those who scored greater than or equal to the mean value of correct responses from 7 item questions prepared to assess comprehensive knowledge of IFAS of the respondents was deemed to have good knowledge about IFAS (13).

### Data collection tools and procedures

A structured, interviewer-administered questionnaire was used to collect data from pregnant women in rural areas of Eastern Tigray. The questionnaire was adapted from previously published studies and national guidelines on IFAS, and it consisted of sections on socio-demographic characteristics, obstetric history, and knowledge of IFAS, service-related factors, and adherence to supplementation. The tool was first prepared in English, translated into Tigrigna, the local language, and then back-translated into English to ensure consistency. Face-to-face interviews were conducted either at health facilities during antenatal care visits or at the respondents’ homes for those who received IFA services from HEWs. The interview approach was chosen to minimize misunderstanding of questions and to include women with limited literacy.

To ensure data quality, daily supervision was carried out, questionnaires were checked for completeness and accuracy at the end of each day, and feedback was provided to data collectors. Completed data were coded and stored securely to maintain confidentiality.

### Data quality assurance

To assure the quality of data, a three-day training was given on the objective of study, data collection process and relevance of the study to both data collectors and supervisors a week before the data collection. Prior to data collection, a pre-test was conducted on 30 pregnant women in Subha Saiesie district area to assess clarity, validity, and reliability of the instrument. Necessary modifications were made based on the findings of the pre-test. The principal investigator was taking corrective measure for any missing values, irregularities, and inconsistencies timely; to ensure data quality at each data collection level.

### Data processing and analysis

After collection the data was imported to SPSS, cleared and analysed using SPSS version 25.0. Categorical variable data were analysed using frequency and percentage, and continuous data was analyzed using measure of central tendency (mean (±SD)). Binary logistic regression model was used. Variables which have P-value of <0.25 were used for analysis in the multivariable logistic regression. Statistical significance was declared at P<0.05 in the multivariable logistic regression model. The goodness of fit the final logistic model was tested by using Hosmer and Lemeshow test at a value of 0.29. Finally, result is presented using texts, figures and tables.

## Results

### Socio-demographic **c**haracteristics of participants

A total of 599 (97.5%) pregnant women participated in the study. The mean (±SD) age of the mothers was 29.1 (± 6.0) years. The majority, 179 (29.9%) were aged between 25 and 29 years. About 275 (45.9%) completed primary education, while 33 (5.5%) had no formal education. Majority, 407 (67.9%) of our study participants were unemployed (**Table 1**).

**Table 1:**
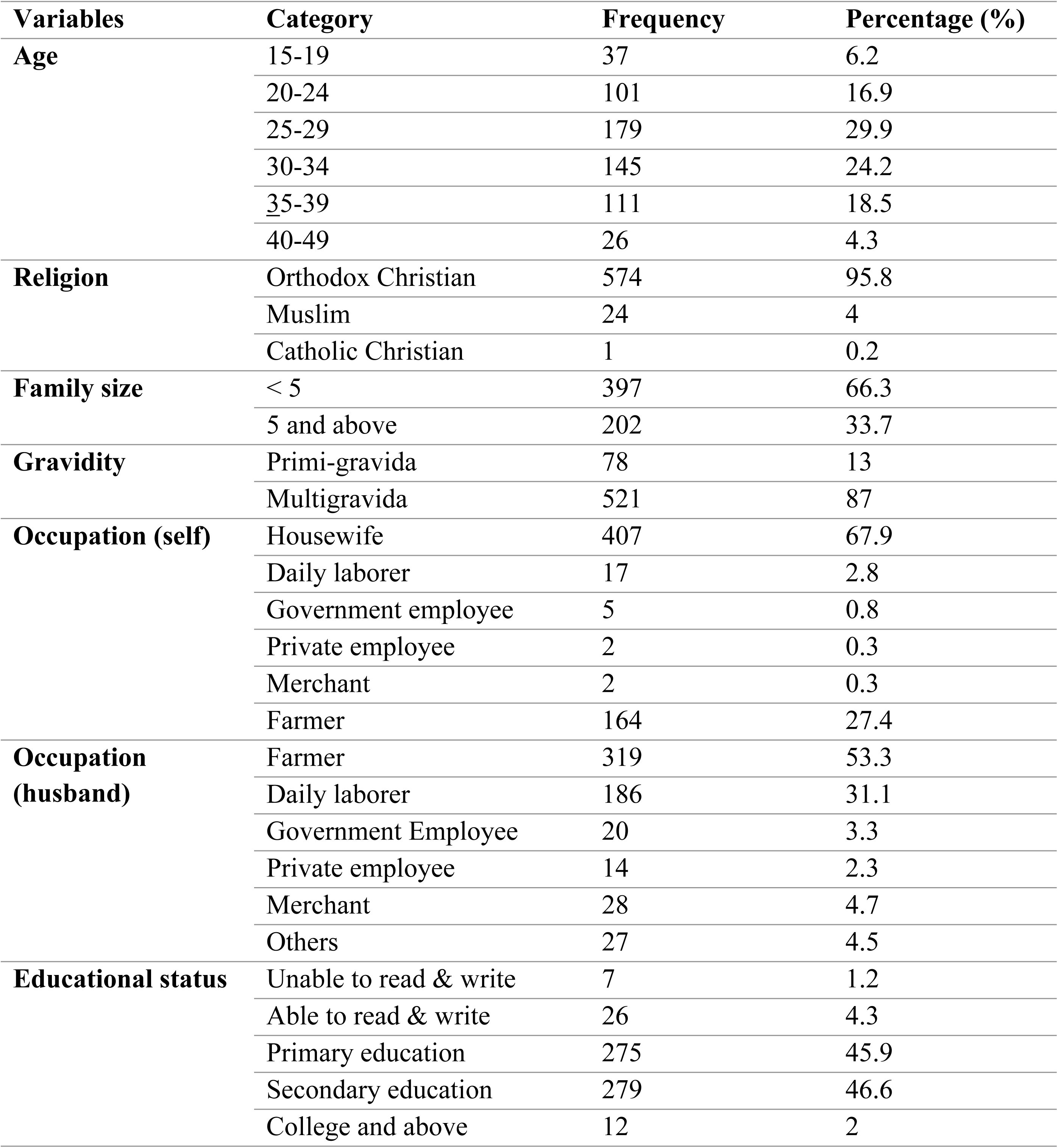
Socio-demographic characteristics among pregnant women in Eastern Tigray, Ethiopia, 2025 (N= 599)

| **Variables** | **Category** | **Frequency** | **Percentage (%)** |
|---|---|---|---|
| **Age** | 15-19 | 37 | 6.2 |
|  | 20-24 | 101 | 16.9 |
|  | 25-29 | 179 | 29.9 |
|  | 30-34 | 145 | 24.2 |
|  | 35-39 | 111 | 18.5 |
|  | 40-49 | 26 | 4.3 |
| **Religion** | Orthodox Christian | 574 | 95.8 |
|  | Muslim | 24 | 4 |
|  | Catholic Christian | 1 | 0.2 |
| **Family size** | < 5 | 397 | 66.3 |
|  | 5 and above | 202 | 33.7 |
| **Gravidity** | Primi-gravida | 78 | 13 |
|  | Multigravida | 521 | 87 |
| **Occupation (self)** | Housewife | 407 | 67.9 |
|  | Daily laborer | 17 | 2.8 |
|  | Government employee | 5 | 0.8 |
|  | Private employee | 2 | 0.3 |
|  | Merchant | 2 | 0.3 |
|  | Farmer | 164 | 27.4 |
| **Occupation (husband)** | Farmer | 319 | 53.3 |
|  | Daily laborer | 186 | 31.1 |
|  | Government Employee | 20 | 3.3 |
|  | Private employee | 14 | 2.3 |
|  | Merchant | 28 | 4.7 |
|  | Others | 27 | 4.5 |
| **Educational status** | Unable to read & write | 7 | 1.2 |
|  | Able to read & write | 26 | 4.3 |
|  | Primary education | 275 | 45.9 |
|  | Secondary education | 279 | 46.6 |
|  | College and above | 12 | 2 |

### Obstetrics and health- related characteristics of participants

From the total respondents, 173 (28.9%) were pregnant for their first time. The median gestational age during first ANC visit was 14.0 weeks. 225 (37.6%) participants visit ANC four times and above (**Table 2**).

**Table 2.** obstetrics and health-related characteristics of pregnant women among pregnant women in Eastern Tigray, Ethiopia, 2025 (N= 599)

| **Variable** | **Category** | **Frequency** | **Percentage** |
|---|---|---|---|
| **Gravidity** | Primigravida | 78 | 13 |
|  | Multigravida | 521 | 87 |
| **Planned pregnancy** | No | 182 | 30.4 |
|  | Yes | 415 | 69.6 |
| **Received counseling about IFAS** | No | 121 | 20.2 |
|  | Yes | 478 | 79.8 |
| **ANC visits frequency** | Less than 4 visits | 374 | 62.4 |
|  | Four visits and more | 225 | 37.6 |
| **History of abortion** | No | 513 | 85.6 |
|  | Yes | 86 | 14.4 |

### Adherence to IFAS

This study revealed the overall adherence to IFAS was 56 % (95% CI: 52% - 60%). The decreased adherence to IFA might be due to lack of knowledge, perceived side effects, were due to side effects, lack of access, and recall bias (**Figure 2**).

**Figure 2.**
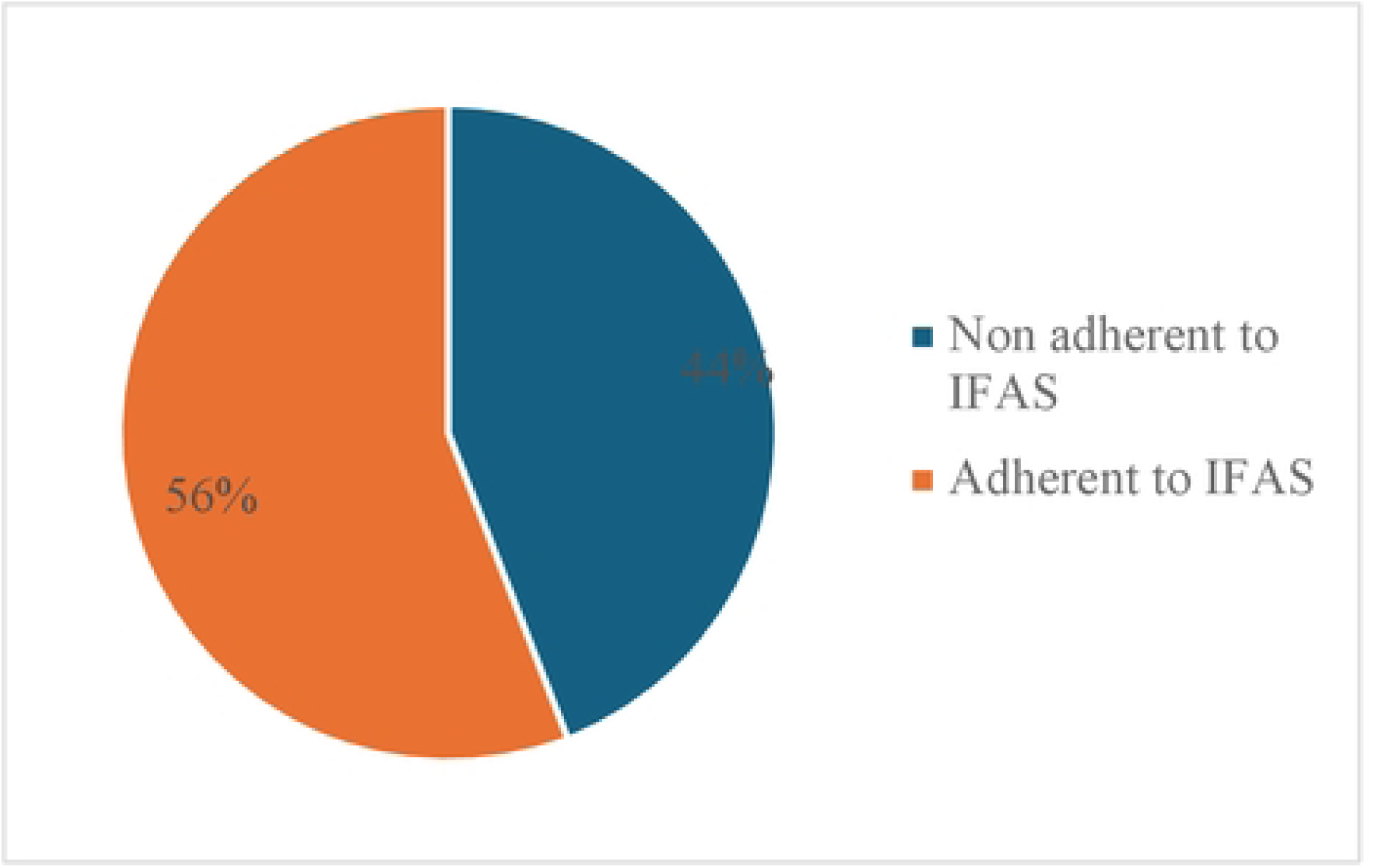
Adherence to IFAS among pregnant women in Eastern Tigray, Ethiopia, 2025.

### Factors associated with adherence to IFAS

After adjusting for potential confounders, pregnant women with four or more ANC visits (AOR= 37.7 95% CI: 14.08-101.03), mothers with previous history of anemia (AOR, 2.2; CI 1.07, 4.69), age of mothers ≥ 40 years (AOR, 11.5; CI: 1.06, 124.05), who received counseling about IFAS (AOR, 7.1; CI: 3.24, 15.37), and mothers travelled to the health facility 30 to 60 minutes and more than 60 minutes ((AOR, 0.02; CI: 0.01, 0.04), and (AOR, 0.01(0.001, 0.04), respectively had statistically significant association with adherence to IFAS (**Table 3**).

**Table 3:** Association of participants’ characteristics with adherence to IFAS among pregnant women in Eastern Tigray, Northern Ethiopia, 2025 (N=559)

| **Variables** | **Categories** | **Adherence to IFAS** |  |  |  |  |
|---|---|---|---|---|---|---|
|  |  | Poor (%) | Good (%) | COR (95% CI) | AOR (95% CI) | P |
| **Age** | 15-19 | 17 (45.9) | 20 (54.1) | 1 | 1 |  |
|  | 20-24 | 42 (41.6) | 59 (58.4) | 1.19 (0.56, 2.55) | 2.2 (0.43, 10.80) |  |
|  | 25-29 | 83 (46.4) | 96 (53.6) | 0.98 (0.48, 2.00) | 1.3 (0.29, 5.58) |  |
|  | 30-34 | 58 (40) | 87 (60) | 1.28 (0.62, 2.64) | 2.2 (0.46, 10.45) |  |
|  | 35-39 | 50 (45) | 61(55) | 1.04 (0.49, 2.19) | 1.7 (0.33, 8.35) |  |
|  | 40-49 | 14 (53.8) | 12 (46.2) | 0.73 (0.27, 1.99) | 11.5 (1.06, 124.05) | 0.045 |
| **ANC Visits** | <4 ANC visits | 240 (64.2) | 134 (35.8) | 1 | 1 |  |
|  | ≥ 4 ANC visits | 24 (10.7) | 201(89.3) | 15 (9.35, 24.08) | 37.7 (14.08, 101.03 | 0.001 |
| **History of anemia** | No | 154 (45.8) | 182 (54.2) | 1 |  |  |
|  | Yes | 110 (41.8) | 153 (58.2) | 1.18 (0.85, 1.63) | 2.2 (1.07, 4.69) | 0.032 |
| **Counseling about IFAS** | No | 89 (73.6) | 32 (26.4) | 1 | 1 |  |
|  | Yes | 175 (36.6) | 303 (63.4) | 4.82 (3.09, 7.51) | 7.1(3.24, 15.37) | 0.001 |
| **Distance to nearest health facility** | < 30 minutes | 39 (11.2) | 309 (88.8) | 1 | 1 |  |
|  | 30-60 minutes | 98 (81) | 23 (19) | 0.03 (0.02, 0.05) | 0.02 (0.01, 0.04) | 0.001 |
|  | > 60 minutes | 127 (98.4) | 2 (1.6) | 0.002 (0.001, 0.004) | 0.01(0.01, 0.04) | 0.001 |

## Discussion

In Ethiopia, IFA remains a significant public health concern, contributing to worsen maternal and neonatal outcomes. Recognizing this challenge, the present study was conducted to evaluate the level of adherence to IFAS among pregnant women, as well as to identify the factors associated with adherence in the Eastern Tigray, Northern Ethiopia. The current study revealed 56% adherence level. And the factors associated with IFAS adherence were ANC visit, previous history of Anemia, distance to health facility, counseling about IFAS and age of the pregnant women.

This study indicated a low rate of adherence to IFAS. This finding was higher than study in Tigray, which was reported 28.9% (10). It is also higher than a systematic review and met analysis conducted on IFAS adherence conducted in Ethiopia which was 41.38% (12). The current study is also higher than the studies conducted in Kenya 24.5% & 32.7% (14, 15) and Uganda 12% (16).

The finding was however lower than study conducted in Addis Ababa which was 70.1% (17), and Mizan Aman town (18), Ethiopia, respectively. The justifications for the differences are variations in socio-economic status, discrepancy in the access of health care facilities, and dissimilarities in the study periods.

In our study, women with four or more ANC visits were 37.7 times more likely to adhere to IFAS as compared those with less than four ANC visits. ANC visit frequency showed a positive association, likely due to increased opportunities for counseling and monitoring. The results support existing literature suggesting that more frequent contact with health workers improves compliance with prenatal recommendations. This finding was supported by those of studies done in Tigray, Ethiopia, and Indonesia (10, 19), respectively. The possible reason might be due to the number of ANC visits increases the opportunity for getting enough iron folate tablets also increases. In addition, one of the essential nutrition action contact points is pregnancy. Therefore, during ANC visits, counseling about iron folate supplementation and advice to continue taking iron folate tablets is expected from health workers who give ANC services. This is in line with the results of several previous studies conducted in SSA (12, 20, 21).

This study showed that women who had history of anemia during their pregnancy were more likely to adhere compared to women who did not have. This finding was like those of studies done in Ethiopia (11, 22) and Tanzania (23). The reason might be that women who had anemia history took more tablets for fear of side effects, and that health providers gave more emphasis to anemic clients than to non-anemic ones. Women who received counseling about IFAS were 7.1 more likely to adhere as compared with their counterparts. This finding is in line with studies conducted elsewhere.

Despite the presence of mixed findings, older age women were more likely to adhere to IFAS (24, 25) as compared with those young once in our case.

Those women who travelled long distance for accessing health were less likely to adhere to IFAS as compared to those found in the walking distance (less than 30 minutes). Our finding is also in line with a study done in other parts of Ethiopia and India (26, 27), and India (28). This finding reinforces evidence that transportation barriers, long travel distances, and poor road infrastructure remain major challenges to accessing maternal health service in Tigray, especially in rural areas as a result women may be discouraged from initiating ANC or attending follow-up visits, thereby reducing their chances of receiving and consistently using IFAS. This study was a community-based study that addressed many relevant issues but it was not free of limitations. It did not show us the cause and effect due to the cross-sectional nature of the study. The questionnaire was self-reported, which might be subject to recall bias.

## Conclusions

The findings of this study indicate that adherence to iron and folic acid supplementation among pregnant women remains below the recommended level. Women having four or more ANC visits, previous history of anemia, receiving counseling about IFAS, older age were the factors positively associated with adherence of IFAS among pregnant women in Easter rural Tigray, Ethiopia. Women who travelled greater than 60 minutes for health seeking were less likely to adhere to IFAS. Hence it has an implication to design strategic policies and to prevent anemia from inadequate intake of iron and folic acid.

## Recommendations

### Recommendation to regional health bureau

✓ Strengthen capacity building (counselling) to the healthcare workers at maternal and child health unit
✓ Address the inaccessibility issue due to the distance to the health facilities, probably by building health posts and addressing through community and home to home

### Recommendation to district health offices

✓ Follow the performance of the healthcare workers, whether health extension workers are really performing according to the written standard both at health post, home to home, at the community out reaches

### Recommendation to pregnant women

✓ Promoting the importance of attending ANC visits should be key priorities

## Data Availability

The datasets generated and/or analyzed during the current study are conceptualized and summarized in the manuscript. For the sake of confidentiality, raw data is not publicly available. However, the datasets can be shared on reasonable requests from the corresponding author.

## Abbreviations

ANC: Antenatal Care
SPSS: Statically Package for Social Science
WHO: World Health Organization

## Declarations

## Ethics approval and consent to participate

Ethical clearance and approval were obtained from Adigrat University College of Medicine and Health Sciences ethical review board. Support letter was obtained from Tigray Regional Health Bureau to the selected district health offices. Before interviewing of participants, each study participant was adequately informed and realized about the purpose of the study and the importance of their participation to confirm willingness for participation. Also, participants were informed that they have a full right to refuse or discontinue participating at any point of the interview. Additionally, participants were informed that the data will not be given for any one and as it is used for the research purpose only to keep its confidentiality.

## Consent for publication

Not applicable.

## Acknowledgements

We would like to express our deepest gratitude to Adigrat University, department of family health and reproductive health, for creating this opportunity. This thesis wouldn’t have been possible without the guidance and encouragement of my supervisors, data collectors, and study participants. Lastly, we acknowledge the support of friends and family for their understanding and encouragement throughout this endeavor.

## Funding

No fund was obtained for this study.

## Competing interests

No competing interest is declared by the authors.

## Notes

### Competing Interest Statement

The authors have declared no competing interest.

### Author Declarations

Ethical clearance and approval were obtained from Adigrat University College of Medicine and Health Sciences ethical review board.

